# Portable Ultra-Low Field MRI Deep-Learning Algorithms for White Matter Lesion Segmentation Improve Accuracy and Reflect Clinical Disability in Multiple Sclerosis

**DOI:** 10.64898/2026.07.15.26357954

**Authors:** Ashley A. Thommana, Corinne A. Donnay, Gina Norato, María I. Gaitán, Ludovica Griffanti, Govind Nair, Daniel S. Reich, Serhat V. Okar

## Abstract

White matter lesion (WML) identification, assessment, and characterization using magnetic resonance imaging (MRI) are fundamental for diagnosis and monitoring of multiple sclerosis (MS). Portable ultra-low field (pULF) MRI at 64 millitesla (mT) has been shown to visualize WML with at least one dimension greater than 4 mm. An automated WML segmentation tool catered to pULF-MRI can provide standardized and accurate quantitative measurements of WML volume. In this study, we sought to investigate and compare the accuracy of machine-learning (ML) and deep-learning (DL) pULF MRI segmentation tools. Same-day paired pULF (64mT) and high-field (HF, 3T) MRI scans from 84 adults with MS or suspected-MS (mean age±SD: 48±13, 62 females) included T2-FLAIR and T1w images. Reference WML segmentations were manually annotated on pULF T2-FLAIR for all scans, with WML confirmed with registered HF T2-FLAIR. HF reference WML segmentations were created. Four automated segmentation methods were applied to pULF scans: Method for Inter-Modal Segmentation Analysis (MIMoSA), an ML algorithm trained on HF WML masks; WMH-SynthSeg, a convolutional neural network model with flexible segmentation capabilities across field strengths and resolution; nnU-Net, a DL algorithm trained on pULF reference WML masks; and Pseudo-Label Assisted nnU-Net (PLAn), a DL algorithm pre-trained on HF reference WML masks and refined with 64mT reference WML masks. Two models were trained with nnU-Net, one using T2-FLAIR images only (nnU-Net-FL) and one using T1w and T2-FLAIR images (nnU-Net-FL/T1). The same was done with PLAn, creating PLAn-FL and PLAn-FL/T1. The six automated WML segmentation outputs were compared to the manual segmentations to determine Dice Similarity Coefficient (DSC) scores.

Associations of WML volume estimates with clinical measures were investigated.

DSC scores with pULF reference WML masks from PLAn-FL (DSC mean±SD: 0.50±0.24) outperformed MIMoSA (0.24±0.20, p<0.0001), WMH-SynthSeg (0.30±0.18, p<0.0001), nnU-Net-FL (0.41±0.24, p<0.0001), and nnU-Net-FL/T1 (0.41 ± 0.26, p = 0.0004). Worse Expanded Disability Status Scale (EDSS) and Scripps Neurologic Rating Scale (SNRS) scores were correlated with higher WML volumes in the pULF and HF reference masks. They were also correlated with WML volumes derived from WHM-SynthSeg, nnU-Net-FL, nnU-Net-FL/T1, PLAn-FL, and PLAn-FL/T1, but not MIMoSA. After adjusting for age, WHM-SynthSeg, nnU-Net FL, nnU-Net-FL/T1, PLAn-FL, and PLAn-FL/T1 had significant associations with EDSS and SNRS scores.

nnU-Net and PLAn performed best in segmenting WML on pULF-MRI at 64 mT, providing accurate quantitative estimates of WML burden. Moreover, WML volumes estimated by these algorithms were associated with clinical measures of disability, underscoring their utility for reflecting clinical and radiological disease severity. Given pULF-MRI’s mobility and lower cost, these findings highlight its relevance in clinical trials, particularly in involving more participants who face logistical constraints and barriers.

**Highlights:**

- Deep learning algorithms were trained on 20 paired, same-day pULF and HF MRI
- Automated pULF segmentation tools were evaluated using 84 paired, same day MRI
- Automated pULF WML segmentations align with reference pULF and HF segmentations
- Trained deep learning algorithms demonstrate strong pULF MRI lesion segmentation
- Automated pULF WML estimates are associated with clinical disability scores

## 1. Introduction

Magnetic resonance imaging (MRI) has been central in the identification of demyelinating white matter lesions (WMLs) in MS, which are essential for diagnosis and disease monitoring^1, 2^. While the pathology of MS neuroinflammation is diverse^3^, the monitoring of WML changes through MRI guides management and may serve as an outcome measure for interventions^4^.

High-field (HF) MRI scanners (>1.5 tesla [T]) are generally implemented to visualize WML in clinical settings^5^, as increasing the magnetic field increases signal-to-noise ratio (SNR), contrast-to-noise ratio (CNR) in some sequences, and achievable resolution^6, 7^. However, HF-MRI infrastructure and maintenance can contribute to the high cost of MRI, which can result in inadequate access, especially in low-resource settings^8^. Portable ultra-low-field (pULF) MRI (at 64 millitesla, mT) is proposed to be more cost-effective^9^, can be transported to bedside to reduce patient burden, and may be safe for some patients with common HF-MRI contraindications^10^. A previous study demonstrated that pULF- MRI consistently captures 3T-confirmed MS lesions larger than 4 mm in diameter^5^. Thus, pULF-MRI has potential implications in MS clinical and research environments, such as triage and clinical trials^10–12^. In such settings, systematic quantification of WML burden on pULF-MRI can enhance its utility.

In contrast to HF- MRI^13–15^, fewer segmentation options exist for pULF-MRI images. At pULF, heterogeneous magnetic field properties, lower SNR, and artifacts resembling WMLs complicate lesion detection^11^, making it challenging to apply HF segmentation models directly. For instance, Method for Inter-Modal Segmentation Analysis (MIMoSA)^14^, a 3T WML segmentation algorithm that uses intermodal coupling regression and structural modeling,^14^ has been applied to same-day paired HF- and pULF-MRI images, demonstrating strong correlation between WML volumes despite a low overlap^5^. The low overlap may reflect challenges arising from higher false positivity due to peripheral artifacts and flow-related venous hyperintensities seen at pULF^5^.

Several other approaches have been used to improve pULF segmentation, including super-resolution methods^16–18^. WMH-SynthSeg^19^, built on SynthSR, a super-resolution deep-learning (DL) model that synthesizes high-resolution isotropic T1-weighted (T1w) images from low-resolution data^16–18^, was designed to segment WML and other brain structures from scans with varying resolutions and contrasts, including pULF-MRI sequences.^17^ In a relatively small cohort of people with MS (n = 12), there was a high correlation of WML volumes with 3T. However, this analysis was limited to volumetric correlations and did not assess segmentation accuracy^19^.

These previous studies evaluated pULF WML segmentation using HF-derived reference masks rather than a pULF-specific ground truth ^5,19^. Given the inherent differences in imaging capabilities between pULF and HF MRI, such as lower spatial resolution and reduced sensitivity to small lesions, the correspondence between pULF segmentations and HF reference masks may be influenced by detectability constraints specific to pULF. Characterizing which lesions are consistently identifiable on pULF relative to paired HF scans, and establishing pULF-specific reference standards, would therefore facilitate more interpretable and realistic evaluation of automated segmentation at this field strength.

DL segmentation models provide a promising avenue for automating lesion segmentation at pULF. Indeed, convolutional neural networks (CNNs) have been widely adopted for MRI segmentation. One example is the nnU-Net method^20^, an open-source 3D biomedical image segmentation tool, which has been shown to be robust in for brain and WML segmentation in HF-MRI^21, 22^. Furthermore, transfer learning can improve WML segmentation performance when data availability is limited by reusing an already trained model as a starting point for a new, related task. We previously demonstrated that Pseudo-Label assisted nnU-Net (PLAn), a transfer learning framework in which an nnU-Net model was pretrained using automated 3T “pseudo-labels” and subsequently fine-tuned on manually annotated 7T data, improved WML segmentation performance at 7T^23^.

In this study, we trained nnU-Net and PLAn models to explore their performance in comparison to other currently available automated segmentation tools and reference segmentations at pULF and HF. We further investigated associations between automated WML volume estimates from pULF-MRI and clinical measures of disability.

## 2. Methods

### 2.1 Participants

Clinical, demographic, and imaging data were acquired from adults with known or suspected MS who underwent same-day pULF (64mT) and HF (3T) MRI scans between April 2021 and June 2024 and had a clinical evaluation within a year (n = 104). Study participants provided written, informed consent under an institutional review board-approved protocol: the National Institute of Neurological Disorders and Stroke’s “Evaluation of Progression in Multiple Sclerosis by Magnetic Resonance Imaging” protocol (NCT00001248). In this protocol, participants with MS undergo annual HF-MRI of the brain with a variety of advanced research pulse sequences, with the option to additionally undergo pULF- MRI; those who consented to the additional scan were recruited for this study.

### 2.2 Clinical & Demographic Data

Demographic data were extracted for all participants. Clinical and radiological presentation were evaluated to categorize participants as MS^24^, other inflammatory neurological disease (OIND), or non-inflammatory neurological disease (NIND). MS participants were further categorized as relapsing-remitting MS (RRMS)^24^, primary progressive MS (PPMS)^24^, secondary progressive MS (SPMS)^24^, clinically isolated syndrome (CIS)^24^, or radiologically isolated syndrome (RIS)^25^. Expanded Disability Status Scale (EDSS) scores^26^, Scripps Neurologic Rating Scale (SNRS) scores^27^, 9-hole peg test (9- HPT)^28^ times from the dominant hand, Paced Auditory Serial Addition Test (PASAT)^29^ score, and Symbol Digit Modalities Test (SDMT)^30^ scores were documented.

### 2.3 MRI Acquisition

HF-MRI was acquired on a 3T Siemens Skyra scanner (software version: VE11C) with a 20- or 32- channel head coil, using a standard protocol including T1w-MP2RAGE and T2-FLAIR scans. pULF- MRI was acquired on portable 64mT SWOOP MRI system (Hyperfine, Guilford, CT) with anisotropic axial T1w and T2-FLAIR sequences Software versions reported in **Supplementary Materials**. MRI sequence parameters are detailed in **Table 1**.

**Table 1:** MRI acquisition parameters for paired HF (3T) and pULF (64mT) scans. *\* All pULF acquisition parameters are provided in Supplemental* Table 1.

| Magnetic Field Strength | Sequence | Acquisition Plane | Repetition Time (s) | Echo Time (s) | Inversion Time (s) | Flip angle (degree) | Slice thickness (mm) | In Plane Voxel Size (mm) | Scan time (min:s) |
| --- | --- | --- | --- | --- | --- | --- | --- | --- | --- |
| *64mT | T2-FLAIR | Axial (3D) | 3.5-4 | 0.16-0.21 | 1.25-1.43 | 90 | 5 | (1.7 x 1.7) – (2 x 2) | 11:19 |
| *64mT | T1-weighted | Axial (3D) | 0.88 – 1.5 | 0.005 - 0.006 | 0.27 - 0.35 | 90 | 5-6 | (1.6 x 1.6) | 6:24 |
| 3T | T1w-MP2RAGE | Sagittal (3D) | 5 | 0.0029 | 0.7/0.25 | 0 | 1 | 1 | 5:38 |
| 3T | T2-FLAIR | Sagittal (3D) | 4.8 | 0.352 | 1.8 | 120 | 1 | 1 | 7:12 |

### 2.4 Datasets for Training and Evaluation

For all participants, reference WML segmentations were created at HF and pULF. HF reference WML segmentations were produced using the Classification using DErived-based Features (C-DEF) segmentation algorithm^13^. AAT (a neuroimaging researcher trained by SVO, a neurologist with seven years of experience in MS neuroimaging) manually refined C-DEF WML masks as needed.

To create pULF reference WML segmentations, two raters (AAT, SVO) followed a consensus approach to manually segment WML visible on the native pULF T2-FLAIR images. WML were confirmed as true-positive by cross-referencing with HF T2-FLAIR rigidly registered to the pULF images^31^ and reformatted to the axial plane.

The paired HF- and pULF-MRI scans, along with their respective reference WML masks, were split into a training (n = 20) and testing (n = 84) dataset. The training dataset included seven mild, seven moderate, and six severe WML burden cases, based on qualitative characterization (as previously described^11^) at the time of clinical reading. Each set was randomly chosen within each WML burden category.

All HF and pULF scans were N4-bias field corrected^10^ and skull-stripped using SynthStrip^11^. The skull-stripped pULF T1w images were rigidly registered to the pULF T2-FLAIR with linear interpolation using the ANTs toolbox^32^. The resulting registration matrix was then applied to the non-skull-stripped images.

### 2.5 Automated Lesion Segmentation Methods

Six automatic lesion segmentation algorithms were applied to the pULF testing cohort: MIMoSA^14^, WMH-SynthSeg^19^, nnU-Net-FL, nnU-Net-FL/T1w, PLAn-FL and PLAn-FL/T1w. **Table 2** summarizes the key characteristics of segmentation algorithms compared in this study.

**Table 2:** Overview of automatic segmentation algorithms used for pULF WML segmentation. Legend: FL = T2-FLAIR; T1w = T1-weighted, WML = white matter lesion, pULF = portable ultra-low field, HF = high field

| Algorithm | Description | Training | Input | Output |
| --- | --- | --- | --- | --- |
| MIMoSA | Voxel-based weighted linear regression | Previously trained model for HF | pULF T2-FLAIR, T1w | WML probability map |
| WMH-SynthSeg | Convolutional neural network (U-Net) trained with synthesized MRI | Previously trained with synthetic data sampled from a generative model | pULF T2-FLAIR | Binarized WML mask with 1x1x1mm<sup>3</sup> resolution |
| nnU-Net-FL | Convolutional neural network (U-Net) with 5-fold cross-validation | Trained on 20 pULF scans and reference WML segmentation masks | pULF T2-FLAIR | Binarized WML mask, SoftMax probabilities |
| nnU-Net-FL/T1w |  |  | pULF T2-FLAIR, T1w | Binarized WML mask, SoftMax probabilities |
| PLAn-FL | Convolutional neural network (U-Net) with 5-fold cross-validation; with refinement | Pre-trained on 20 semi-automated WML segmentations, created by correcting HF WML masks output by C-DEF. Model refined with on 20 manual WML segmentation masks at pULF. | pULF T2-FLAIR | Binarized WML mask, SoftMax probabilities |
| PLAn-FL/T1w |  |  | pULF T2-FLAIR, T1w | Binarized WML mask SoftMax probabilities |

For MIMoSA and WMH-SynthSeg, pretrained models were applied to the pULF testing cohort without retraining. MIMoSA was executed on normalized pULF FLAIR and registered T1w images to generate voxelwise probability maps, which were thresholded to produce binary WML masks. WMH-SynthSeg was applied to preprocessed pULF FLAIR images using FreeSurfer’s implementation, and the WML label was extracted and resampled to native pULF resolution. Details of MIMoSA and WMH-SynthSeg implementation on the pULF testing cohort are provided in the **Supplementary Materials**.

The two nnU-Net segmentation models were trained on the pULF training dataset using the publicly available nnU-Net^20^ package (https://github.com/MIC-DKFZ/nnUNet). The first nnU-Net model was trained using only the preprocessed pULF T2-FLAIR (referred to as “nnU-Net-FL”) and the second using the preprocessed registered pULF T1w and pULF T2-FLAIR (referred to as “nnU-Net-FL/T1w”). Both models were trained using 5-fold cross-validation and the 3d_cascade_fullres configuration^20^.

Two PLAn models were trained using the paired HF and pULF training dataset. The PLAn models were pretrained using HF scans and HF reference WML masks, both downsampled to 1.7x1.7x5 mm. Following the methods in our previous work^23^, transfer learning was performed by loading the weights of the pretrained model and the corresponding processing parameters (excluding the final softmax layer). The model was then fine-tuned using the pULF training dataset. One PLAn model used only T2-FLAIR images (PLAn-FL), and the other using T2-FLAIR images and registered T1w images (PLAn-FL/T1w).

### 2.6 Quantitative Analysis of Reference WML Segmentations

To establish what WML segmentation is achievable at pULF, total lesion volume agreement and segmentation overlap between pULF and HF reference WML masks were evaluated. To calculate overlap metrics, pULF scans and their reference WML masks were up-sampled to match the voxel size of the HF scans (1mm isotropic). The skull-stripped HF T2-FLAIR scans were rigidly registered to the up-sampled skull-stripped pULF T2-FLAIR using Advanced Normalized Tools (ANTs)^32^, and the registration matrix was applied to the HF reference WML masks. To minimize resampling errors and preserve the original lesion volume, all resampled WML masks were linearly interpolated and thresholded at a value that minimized the difference in volume relative to the original lesion mask volume, selected using iterative thresholds in steps of 0.05.

Individual lesions from five cases were visually evaluated to determine which were captured by the pULF reference WML masks. Lesions in the HF reference masks were classified as “pULF-visible” if present in the pULF reference mask, or “pULF-missed” if absent. Cases with large confluent lesions were excluded to allow characterization of distinct WML. For each lesion, volume and topographic category were extracted.

### 2.7 Quantitative Analysis of Automated Lesion Segmentation Methods

WML segmentation algorithms were evaluated with respect to pULF reference masks. Volumetric and lesion count agreement between reference and automatic WML masks were calculated from the original output of each segmentation method. Overlap metrics were extracted with FSL’s *bianca_overlap_tools*^15^ relative to the pULF reference WML in native pULF acquisition resolution.

Lesion-wise metrics and lesion counts were derived by identifying connected components, where each contiguous cluster of non-zero voxels was treated as an individual lesion. Additional quantitative analyses, including overlap measures calculated in 1×1×1 mm³ space and assessment of optimal thresholding for probability-based segmentation outputs, are provided in the **Supplementary Materials**.

### 2.8 Qualitative Analysis of Automated Lesion Segmentation Methods

In a subset of 24 participants, a blinded rater (SVO, a neurologist with 4 years of experience in pULF imaging) assessed the quality of pULF WML segmentations. 8 participants were randomly selected within each lesion burden group, and anonymized labels were assigned to each segmentation method. A registered HF-T2 FLAIR was provided for reference. The ratings were based on a scale from 1 to 5 as defined in **Table 3**.

**Table 3:** Qualitative scoring scale for pULF segmentation methods. Legend: pULF-MRI = portable ultra-low field MRI; WML = white matter lesion

| Score | Rating | Definition |
| --- | --- | --- |
| 1 | Limited | 0-25% of WML visible on pULF-MRI segmented, 100-75% not segmented or other structures segmented as WML |
| 2 | Modest | 25-50% of WML visible on pULF-MRI segmented, 75-50% not segmented or other structures segmented as WML |
| 3 | Good | 50-75% of WML visible on pULF-MRI segmented, 50-25% not segmented or other structures segmented as WML |
| 4 | Very Good | 75-100% of WML visible on pULF-MRI segmented, 25-0% not segmented or other structures segmented as WML.<br>*In the absence of pULF visible WML, minimal false positive identification |
| 5 | Excellent | Almost all lesions visible on pULF-MRI segmented, well defined and matching the shape of the lesions, minimal false positive areas |

### 2.9 Statistical Analyses

Statistical analyses were conducted in GraphPad Prism 10.1.0. Data were checked for normality with the D’Agostino & Pearson test. Differences between categorical and continuous variables between the training and testing cohort were performed with two-tailed Fisher’s Exact and Mann Whitney U tests, respectively.

Agreement between pULF and HF reference WML volumes was assessed with Spearman correlation and the Wilcoxon signed-rank test. Bland-Altman analysis was used to determine volumetric bias (mean HF - pULF volume) and limits of agreement (bias ± 1.96 × SD).

Volumetric and lesion count agreement between automated segmentation methods and the pULF reference were assessed with Spearman correlation. Lesion-wise detection metrics (true positive, false positive, and false negative lesions) were derived from connected component analysis. Spatial overlap was assessed using the Dice Similarity Coefficient (DSC), with lesion-wise and voxel-wise precision and sensitivity reported separately. Differences across segmentation methods were compared with a Friedman test followed by Dunn’s post-hoc correction. Unadjusted and age-adjusted associations between segmentation volumes and clinical or imaging measures were assessed with linear regression models fitted in R. This work utilized the computational resources of the NIH HPC Biowulf cluster (https://hpc.nih.gov).

## 3. Results

### 3.1 Demographic and clinical characteristics

A total of 104 adults with known or suspected MS (mean age ± SD = 50 ± 13, female = 77, median EDSS = 2.0) were included in the training and testing cohorts. Details for demographic and clinical characteristics of different cohorts are provided in **Table 4**. The median difference between clinical evaluation and imaging was 12 days (range: 0 – 336 days). There were no differences in clinical characteristics between the training and testing cohorts. Notably, reference WML segmentations volumes at pULF and HF were comparable.

**Table 4:**
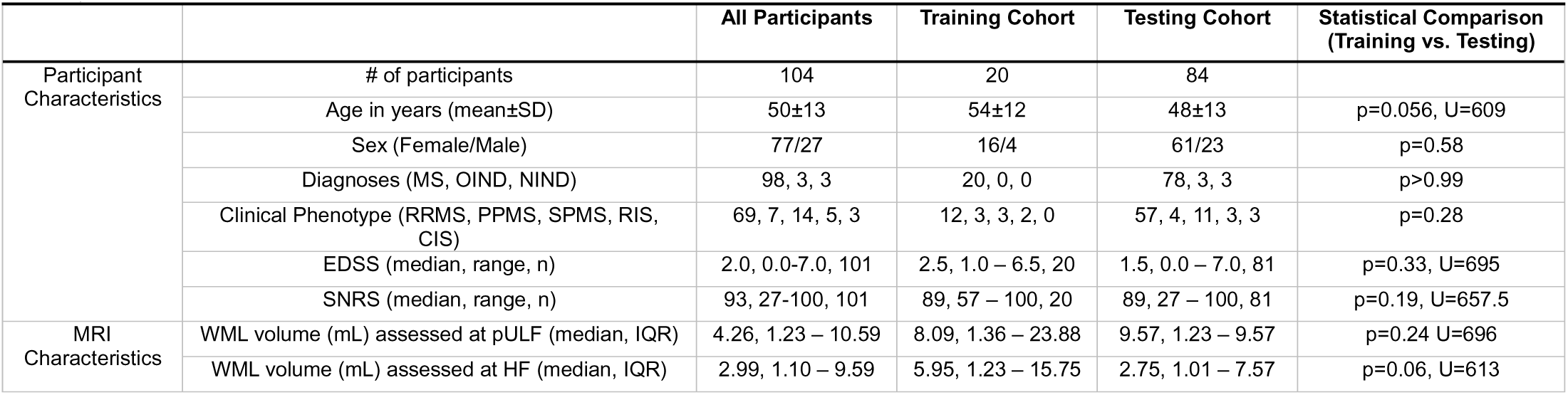
Cohort characteristics and descriptive statistics for all participants (n=104). Abbreviations: MS = multiple sclerosis; OIND = other inflammatory neurological disease; NIND = non-inflammatory neurological disease; RRMS = relapsing remitting MS; SPMS = secondary progressive MS; PPMS = primary progressive MS; EDSS = expanded disability status scale; SNRS = Scripps Neurologic Rating Scale, WML = white matter lesion; pULF = portable ultra-low field; HF = high field. *Continuous variables: Mann–Whitney U test (two-tailed; U statistic and p-value shown). Categorical variables: Fischer’s Exact test (two-tailed for 2 categories; p-value
640 shown)

### 3.2 pULF reference WML segmentation volumes are concordant with HF reference WML segmentation volumes

Total reference WML volumes at pULF strongly correlated with HF reference segmentations (ρ(82) = 0.96, p < 0.0001). However, total lesion volume was higher at pULF compared to HF, with a median difference of 0.82mL (p < 0.0001, W = 2464, Bland-Altman: bias ± SD=1.7 ± 3.21mL, limits of agreement = -4.5 – 8.02mL). The median DSC overlap between reference segmentations was 0.32 (IQR: 0.17 – 0.43).

For lesion-level analyses, 126 distinct lesions were identified at HF across 5 cases. 51 (40%) were captured by the pULF reference WML segmentations. 58 lesions at HF had a volume greater than 50µL (approximating a sphere of diameter ∼ 4.5 mm). pULF reference WML segmentations captured 46 (78%) of these 59 lesions. Of the 66 WML that were smaller than 50µL, only 5 (7%) were captured in the pULF reference WML segmentations. pULF WML reference segmentations captured periventricular lesions best, visualizing 40 of 64 (62.5%) HF lesions. 8 of 41 (19.5%) deep white matter and 3 of 16 (19%) juxtacortical HF lesions were captured by the pULF reference segmentations. 2 callosal, 2 cerebellar, and 1 brain stem HF lesions were not well visualized at pULF and thus were not included on pULF reference masks. Lesion-level performances of segmentation models are reported in the **Supplementary Methods**.

**Figure 1** showcases representative pULF and HF reference WML segmentations (A) and WML volume associations (B, C) across the test cohort. Lesion-level capture rates and topographic distribution from the subset of 126 lesions are shown in panels D and E.

**Figure 1:**
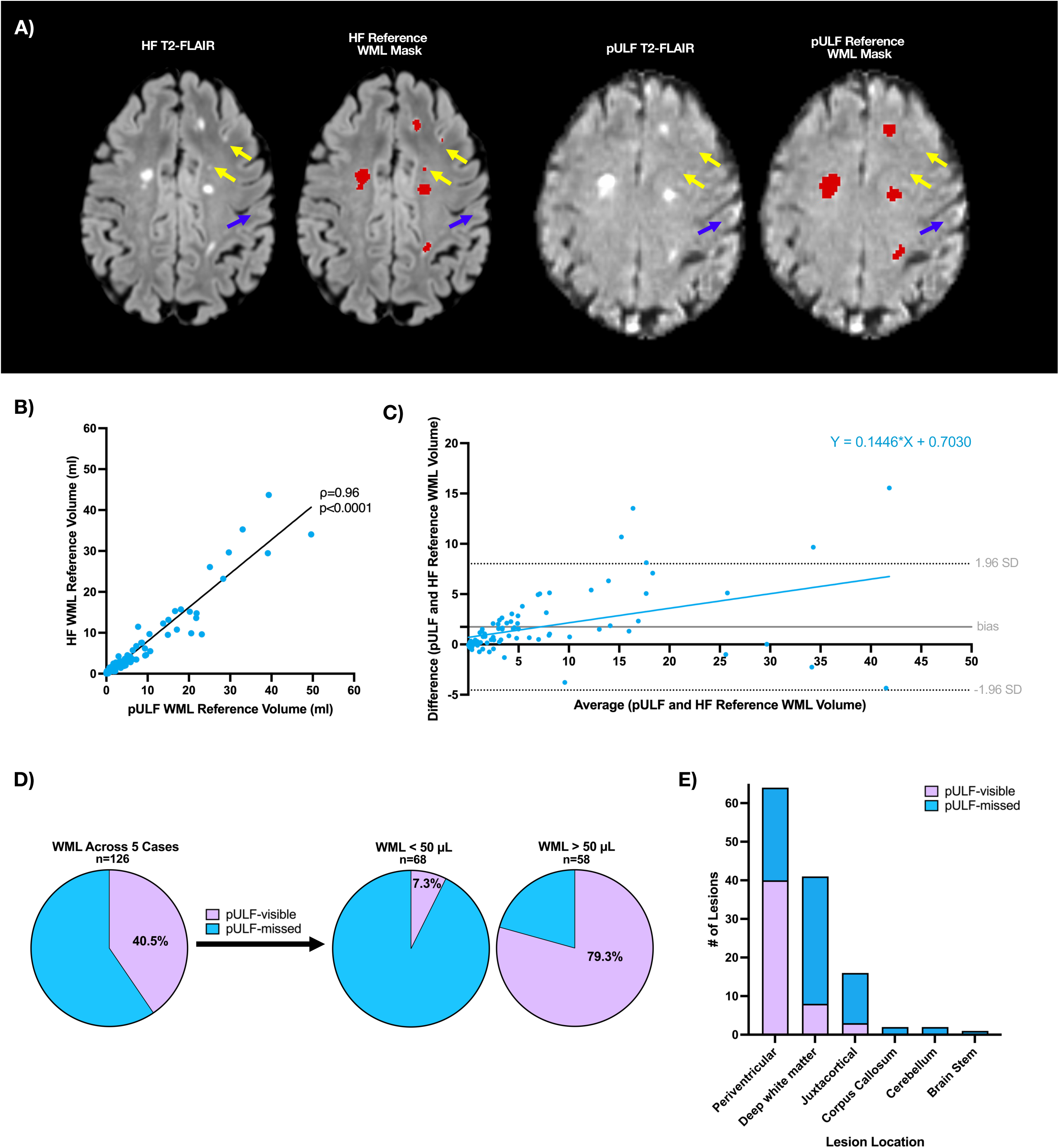
pULF MRI visualizes many lesions seen at HF. A) Representative reference WML masks at HF and pULF for a woman in her 60s with SPMS highlighting comparable patterns for large lesions, though some small lesions (indicated with yellow arrows) were missed at pULF. Misleading hyperintensities resembling WML (indicated with blue arrows) on pULF-MRI were not included in reference masks after comparison with corresponding areas of HF-MRI that were normal-appearing. B) Across the test cohort, pULF reference WML volumes were strongly correlated with HF reference WML volumes. C) Bland-Altman analysis of the test cohort revealed that pULF overestimated total WML volume, particularly at high lesion burden. D) In a subset of 126 distinct lesions, pULF WML reference segmentations captured 40.4% lesions detected at HF. For lesions above 50µL, 79% were captured in the ULF reference segmentations (even after excluding the large confluent lesions), while only 7% of WML at HF that were smaller than 50µL were seen at pULF. E) At the lesion-level within the same subset, pULF reference segmentations captured 62.5% of periventricular, 19.5% of deep white matter, and 18.8% of juxtacortical WML seen at HF.

### 3.3 PLAn segmentation models outperform other segmentation methods in quantitative and qualitative performance metrics

Quantitative performance metrics are summarized in **Table 5**. WML volumes across all automated segmentation methods were correlated (p < 0.0001) with the pULF reference WML segmentation volumes. PLAn models exhibited the strongest volume correlations (PLAn-FL: ρ(82) = 0.90, PLAn- FL/T1w: ρ(82) = 0.90), followed by nnU-Net-FL/T1w (ρ(82) = 0.83), nnU-Net-FL (ρ(82) = 0.80), WMH-SynthSeg (ρ(82) = 0.69), and MIMoSA (ρ(82) = 0.41). Lesion count correlations with the pULF reference were significant for PLAn and nnU-Net models (PLAn-FL: ρ = 0.68, PLAn-FL/T1w: ρ = 0.69; nnU-Net-FL: ρ = 0.53, nnU-Net-FL/T1w: ρ = 0.59; all p < 0.0001), but not for MIMoSA (ρ = 0.054) or WMH-SynthSeg (ρ = 0.039), both of which substantially overestimated lesion count.

**Table 5.** Quantitative performance metrics for automated white matter lesion segmentation methods compared to the pULF reference standard. Metrics are reported as median [interquartile range] unless otherwise indicated, across 84 participants (lesion overlap metrics computed in the 80 participants with at least one WML in the pULF reference mask). Lesion Burden Agreement includes total WML volume (µL), lesion cluster count, and Spearman correlations (ρ) of total WML volume and lesion count with the pULF reference. Lesion Detection metrics are lesion-wise, where individual connected components are treated as discrete lesions; precision reflects the proportion of detected lesions that are true positives, and sensitivity reflects the proportion of reference lesions detected. Spatial Overlap metrics are voxel-wise; the Dice Similarity Coefficient (DSC) reflects overall overlap, while voxel precision and sensitivity reflect the balance of false positive and false negative voxels, respectively. WML: white matter lesion; IQR: interquartile range; FL: FLAIR; T1w: T1-weighted; pULF: portable ultra-low field MRI; DSC: Dice Similarity Coefficient.

|  | MIMoSA | WMH-SynthSeg | nnU-Net (FL) | nnU-Net (FL/T1w) | PLAn (FL) | PLAn (FL/T1w) |
| --- | --- | --- | --- | --- | --- | --- |
| Lesion Burden Agreement |  |  |  |  |  |  |
| Total WML volume in mL (median [IQR]) | 18.93<br>[13.26–25.16] | 7.70<br>[5.69–11.29] | 1.58<br>[0.35–5.31] | 1.26<br>[0.29–5.18] | 2.89<br>[0.83–8.36] | 2.46<br>[0.72–8.27] |
| Lesion clusters (median [IQR]) | 90.50<br>[62.25–118.50] | 16.00<br>[13.50–21.00] | 5.00<br>[2.00–7.00] | 4.50<br>[2.00–6.25] | 6.00<br>[3.00–9.00] | 6.00<br>[3.00–9.00] |
| Total WML volume correlation with pULF Reference (ρ) | 0.41 | 0.70 | 0.80 | 0.83 | 0.90 | 0.90 |
| Total lesion count correlation with pULF Reference (ρ) | 0.054 | 0.039 | 0.53 | 0.59 | 0.68 | 0.69 |
| Lesion Detection (lesion-wise) |  |  |  |  |  |  |
| True positive lesions (median [IQR]) | 7.00<br>[3.00–11.00] | 5.00<br>[3.00–8.25] | 4.00<br>[2.00–7.00] | 4.00<br>[2.00–6.00] | 5.00<br>[2.00–7.00] | 5.00<br>[2.00–8.00] |
| False negative lesions (median [IQR]) | 2.00<br>[1.00–3.00] | 2.00<br>[1.00–5.00] | 3.00<br>[2.00–6.25] | 4.00<br>[2.00–6.00] | 3.00<br>[2.00–5.00] | 3.00<br>[2.00–5.00] |
| False positive lesions (median [IQR]) | 82.00<br>[58.00–117.50] | 10.00<br>[7.00–13.00] | 0.00<br>[0.00–1.00] | 0.00<br>[0.00–1.00] | 1.00<br>[0.00–2.00] | 1.00<br>[0.00–2.00] |
| Lesion precision (median [IQR]) | 0.08<br>[0.03–0.13] | 0.33<br>[0.22–0.50] | 1.00<br>[0.80–1.00] | 1.00<br>[0.88–1.00] | 0.83<br>[0.71–1.00] | 0.85<br>[0.67–1.00] |
| Lesion sensitivity (median [IQR]) | 0.71<br>[0.54–0.81] | 0.67<br>[0.54–0.82] | 0.50<br>[0.25–0.68] | 0.50<br>[0.25–0.67] | 0.60<br>[0.40–0.75] | 0.58<br>[0.40–0.75] |
| Spatial Overlap (voxel-wise) |  |  |  |  |  |  |
| Dice / Similarity Index (median [IQR]) | 0.20<br>[0.05–0.37] | 0.29<br>[0.14–0.44] | 0.43<br>[0.22–0.63] | 0.44<br>[0.20–0.65] | 0.55<br>[0.37–0.68] | 0.53<br>[0.32–0.68] |
| Voxel precision (median [IQR]) | 0.12<br>[0.03–0.32] | 0.22<br>[0.09–0.43] | 0.85<br>[0.69–0.95] | 0.86<br>[0.72–0.95] | 0.71<br>[0.60–0.82] | 0.72<br>[0.59–0.81] |
| Voxel sensitivity (median [IQR]) | 0.46<br>[0.28–0.58] | 0.42<br>[0.30–0.50] | 0.33<br>[0.15–0.54] | 0.31<br>[0.12–0.56] | 0.50<br>[0.27–0.66] | 0.49<br>[0.27–0.65] |

MIMoSA estimated higher total median WML volume (18.93 [IQR 13.26 – 25.16] mL) compared to the pULF reference (4.17 [IQR 1.2 – 9.57] mL, Z = 6.6, p < 0.0001). In contrast, the nnU-Net models (nnU-Net-FL: 1.58 [IQR 0.35 – 5.31] mL; nnU-Net-FL/T1w: 1.26 [IQR 0.29 – 5.18] mL) estimated lower mean WML volumes (Z = 8.3, p < 0.0001; Z = 8.2, p < 0.0001, respectively). Total WML volume from PLAn models (PLAn-FL: 2.89 [IQR 0.83 – 8.36] µL; PLAn-FL/T1w: 2.46 [IQR 0.72 – 8.27] mL) and WMH-SynthSeg (7.7 [IQR 5.69 – 11.29] mL) were comparable with the pULF reference volumes (Z = 2.3, p = 0.65, Z = 2.6, p = 0.24; Z = 2.8, p = 0.16, respectively; **Figure 2A**).

**Figure 2:**
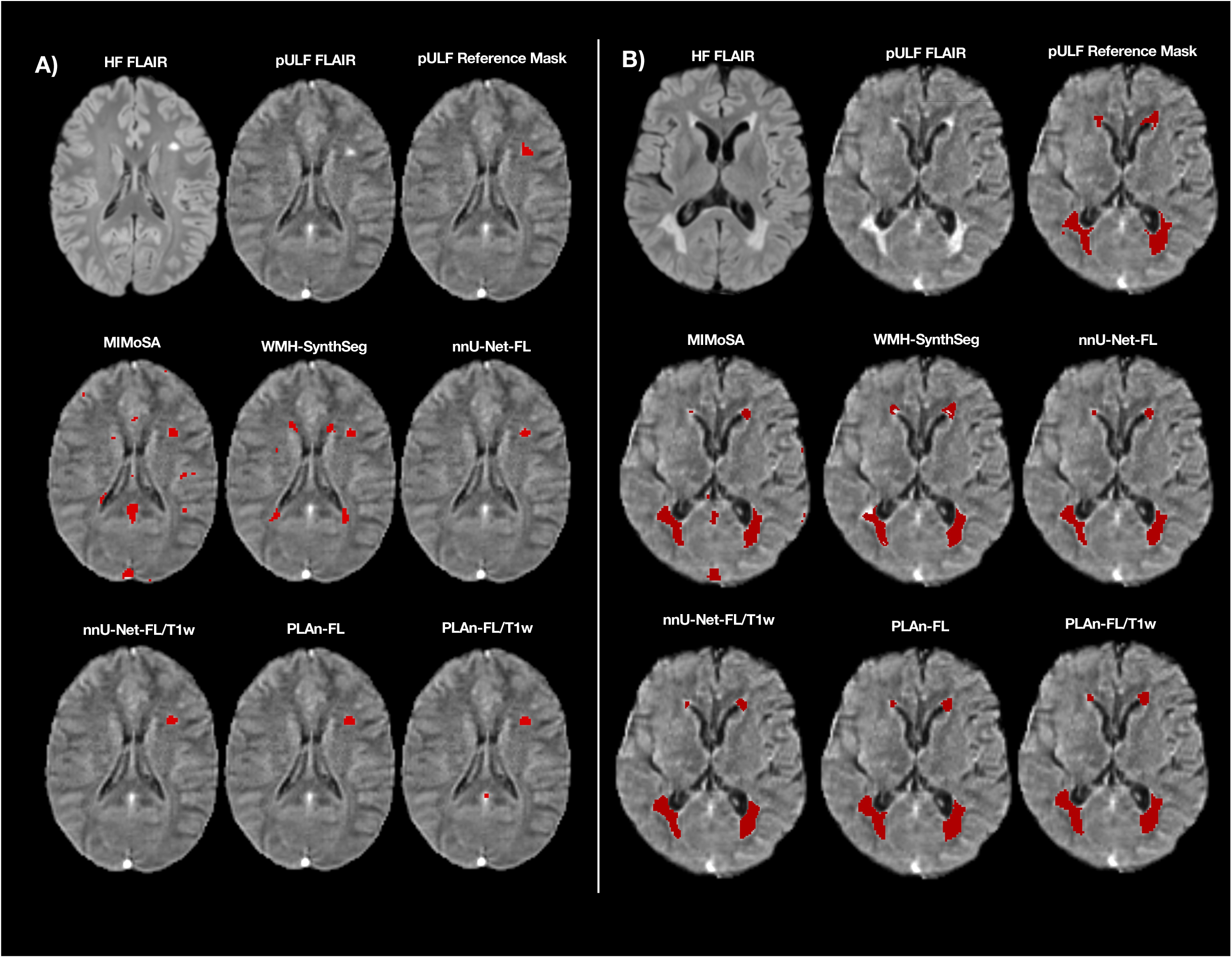
PLAn models demonstrated the highest overlap with pULF reference WML segmentations, with low false positive and negative lesion clusters. A) Total WML volumes from WMH-SynthSeg and PLAn (-FL and -FL/T1w) models were not significantly different from the pULF reference volumes, while MIMoSA overestimated total WML volume and nnU-Net (-FL and -FL/T1w) models underestimated total WML volumes. Pairwise differences were calculated between pULF reference (gray) and automated methods. Significant statistics reported. B) PLAn-FL achieved the highest median DSC among all automatic segmentation methods. Only significant pairwise statistics between PLAn-FL (green) and other methods are shown. C) MIMoSA (blue) and WMH-SynthSeg had the highest false positive lesion counts per scan. All significant pairwise statistics between all methods except for MIMoSA are shown. Pairwise statistics between MIMoSA and WMH-SynthSeg are also shown. D) nnU-Net models had the highest count of false negative lesions per scan. Only significant pairwise statistics between PLAn-FL (green) and other methods are shown. E) MIMoSA (blue) had the highest true positive lesion count, although this may be influenced in part by its tendency towards oversegmentation. nnU-Net models had the lowest true positive counts per scan. All significant pairwise statistics are show. F) In a blinded qualitative rating of segmentation models, PLAn-FL and PLAn-FL/T1 had the highest ratings, evaluated as “Very Good” or “Excellent” in 75% and 87.5%, respectively, of the cases.

In the 80 of 84 participants with at least one WML in the pULF reference mask, PLAn and nnU-Net models (both -FL and -FL/T1 variants) had significantly higher DSC scores than MIMoSA (0.20 [0.05 – 0.37]) and WMH-SynthSeg (0.29 -0.14 – 0.44]). PLAn models achieved the highest performance (PLAn-FL: 0.55 [0.37 – 0.68]; PLAn-FL/T1: 0.53 [0.32 – 0.68]), outperforming nnU-Net models (nnU-Net-FL: 0.43 [0.22 – 0.63], Z=4.0, p < 0.001; nnU-Net-FL/T1: 0.44 [0.2 – 0.65], Z = 3.2, p = 0.002; **Figure 2B**). nnU-Net models were highly conservative (**Figure 2C**), with near-zero false positive lesion counts (median: 0 [IQR 0–1]) and high lesion-wise precision (median: 1.00), but at the cost of reduced sensitivity (median: 0.50 [IQR 0.25–0.68]); this pattern was similarly reflected at the voxel level (voxel precision: 0.85–0.86; voxel sensitivity: 0.31–0.33). PLAn models achieved a more balanced trade-off, with low false positive counts (median: 1 [IQR 0–2]), high lesion precision (median: 0.83–0.85), and better sensitivity (median: 0.58–0.60; voxel sensitivity: 0.49–0.50). In contrast, MIMoSA demonstrated extensive oversegmentation (median false positives: 82 [IQR 58– 118]; lesion precision: 0.08), though this inflated sensitivity at the lesion level (median: 0.71). WMH-SynthSeg also oversegmented, albeit less severely than MIMoSA (median false positives: 10 [IQR 7– 13]; lesion precision: 0.33; sensitivity: 0.67; **Figure 2D**). MIMoSA, however, had the highest true positive lesion counts (median: 7 [IQR 3 – 10.75], **Figure 2E**). This may have been due to its tendency towards oversegmentation. nnU-Net models had the lowest true positive counts (nnU-net- FL median: 4 [IQR 1.25 – 6.75], nnU-net-FL/T1w median: 4 [IQR 1 – 6]). WMH-SynthSeg (median: 5 [IQR 3 – 7.75]), PLAn-FL (median: 5 [IQR 2 – 7]), and PLAn-FL/T1w (median: 4 [IQR 2 – 8]) did not have significantly different true positive counts.

The addition of T1w images as a training input alongside FLAIR did not significantly improve performance (DSC) for either PLAn (PLAn-FL vs PLAn-FL/T1w: Z=0.5, p>0.9) or nnU-Net (nnU-net- FL vs nnU-net-FL/T1w: Z=0.4, p>0.9) models (**Figure 2B**). Neither PLAn nor nnU-Net models showed meaningful improvement in performance with the addition of T1w to FLAIR input, across all quantitative metrics (**Table 5**; **Supplementary Materials**). WMH-SynthSeg’s DSC was consistent across both native and resampled spaces, with no significant differences in overlap trends. All pairwise comparisons for all overlap metrics at down- and up-sampled resolutions are available in the **Supplementary Materials**. Furthermore, fine-tuning of PLAn, nnU-Net, and MIMoSA segmentations using probability thresholds other than the default resulted in stable overlap statistics compared to other methods (reported in the **Supplemental Materials**).

### 3.4 PLAn models were rated highly in qualitative segmentation ratings

Qualitative ratings were consistent with the quantitative findings, with PLAn models receiving the highest scores (PLAn-FL median: 4 [IQR 3.25 – 4.75]; PLAn-FL/T1w median: 4 [IQR 4 – 4.75]; Z = 0.5, p>0.99) and MIMoSA the lowest (1.92 ± 0.15), significantly below all methods except WMH-SynthSeg (median: 3 [IQR 2 – 3];, Z = 2.6, p = 0.15; **Figure 2F**). Representative examples of automatic segmentation outputs in a low and a severe lesion burden cases are shown in **Figure 3A and 3B**, illustrating key patterns observed. nnU-Net models were consistently conservative, both undersegmenting detected lesions and missing lesions that PLAn models correctly identified.

**Figure 3:**
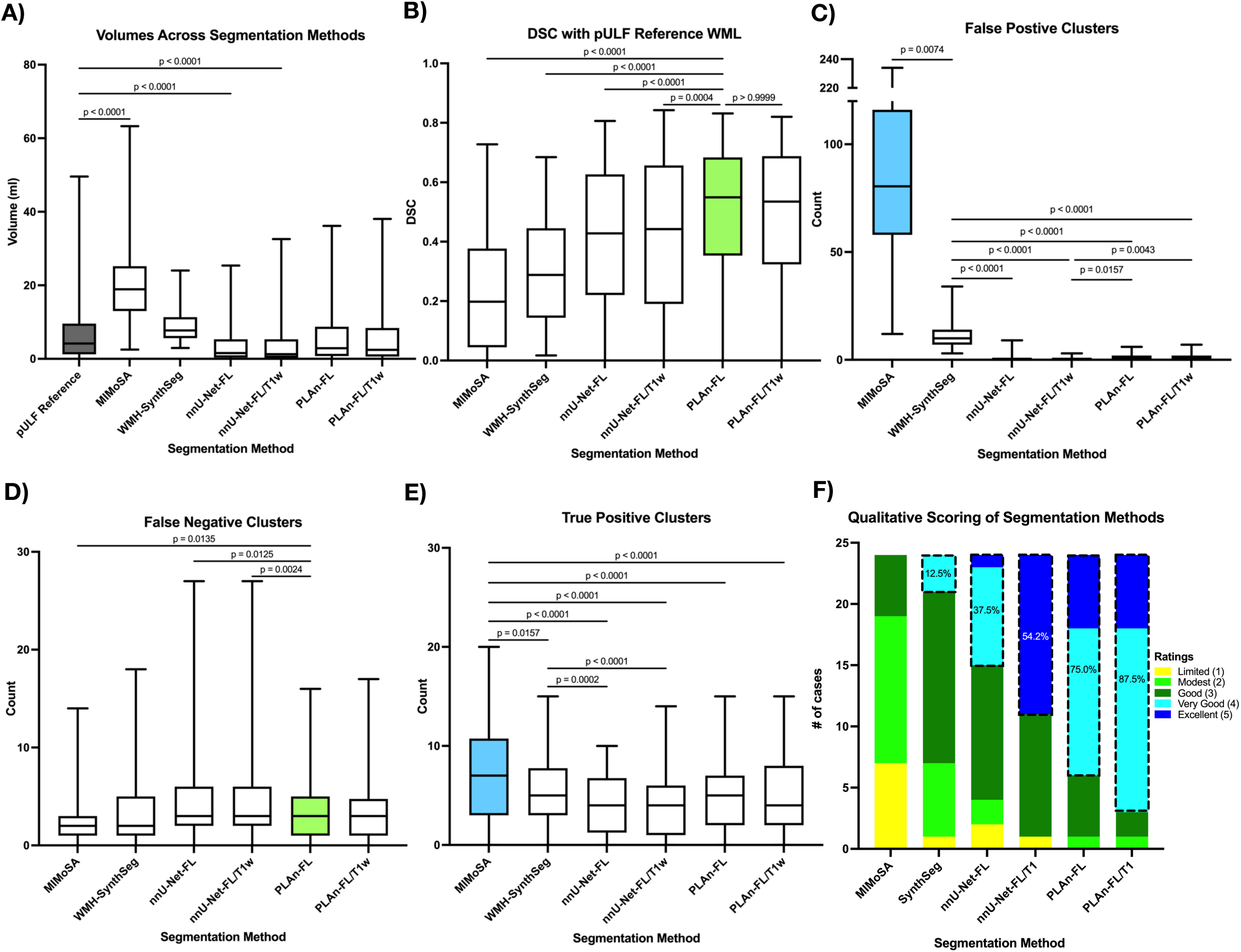
Automated WML segmentations capture lesions in low and high lesion burden cases. A) Representative pULF reference and automatic segmentation WML masks in a woman in her 30s with RRMS and low lesion burden and (B) in a woman in her 40s with RRMS and high lesion burden.

MIMoSA produced extensive false positive detections in hyperintense non-lesion structures, such as the sagittal sinus. WMH-SynthSeg performed better overall, with many errors attributable to oversegmentation of periventricular regions; however, FLAIR hypointense regions were also occasionally mislabeled as lesions.

### 3.5 White matter lesion volumes estimated from automated methods at pULF are associated with age and clinical disability in MS

**Table 6** summarizes significant associations between WML volumes and age and clinical disability scores. In linear models predicting clinical disability scores from WML volume, both HF and pULF reference WML volumes were significantly associated with EDSS (3T: β = 0.08mL^-1^, 95% CI [0.031, 0.129], p = 0.002; pULF: β = 0.068 mL^-1^, 95% CI [0.025, 0.11], p = 0.002) and SNRS disability scores (3T: β = −0.697 mL^-1^, 95% CI [−1.029, −0.365], p < 0.001; pULF: β = −0.652 mL^-1^, 95% CI [−0.934, −0.369], p < 0.001). Similarly, WMH-SynthSeg, nnU-Net (FL and FL/T1w), and PLAn (FL and FL/T1w) segmentation methods demonstrated significant associations with both EDSS and SNRS scores. In contrast, MIMoSA-derived WML volumes were not significantly associated with these clinical measures.

**Table 6.** WML volume and clinical disability associations across automated matter lesion segmentation methods with and without age-adjustment. Fitted multiple linear regression models of WML volume and clinical disability measures (EDSS and SNRS) demonstrated significant associations for both HF and pULF reference segmentations and all automated segmentation models except MIMoSA. Age-adjusted models demonstrated the same findings.

|  | Unadjusted Model |  | Age-Adjusted Model |  |
| --- | --- | --- | --- | --- |
| | $\beta$ (95% CI) in mL<sup>-1</sup> | p-value | $\beta$ (95% CI) in mL<sup>-1</sup> | p-value |
| **EDSS** |  |  |  |  |
| HF Reference | 0.08<br>(0.031, 0.129) | 0.002 | 0.06<br>(0.012, 0.109) | 0.016 |
| pULF Reference | 0.068<br>(0.025, 0.11) | 0.002 | 0.051<br>(0.008, 0.093) | 0.02 |
| MIMoSA | 0.012<br>(-0.025, 0.05) | 0.51 | 0.019<br>(-0.016, 0.054) | 0.282 |
| WMH-SynthSeg | 0.168<br>(0.087, 0.249) | <0.001 | 0.128<br>(0.041, 0.214) | 0.005 |
| nnU-Net-FL | 0.129<br>(0.06, 0.198) | <0.001 | 0.106<br>(0.038, 0.173) | 0.002 |
| nnU-Net-FL/T1w | 0.117<br>(0.061, 0.174) | <0.001 | 0.097<br>(0.041, 0.153) | <0.001 |
| PLAn-FL | 0.096<br>(0.046, 0.146) | <0.001 | 0.076<br>(0.026, 0.126) | 0.003 |
| PLAn-FL/T1w | 0.092<br>(0.045, 0.139) | <0.001 | 0.073<br>(0.026, 0.12) | 0.003 |
| **SNRS** |  |  |  |  |
| HF Reference | -0.697<br>(-1.029, -0.365) | <0.001 | -0.615<br>(-0.955, -0.274) | <0.001 |
| pULF Reference | -0.652<br>(-0.934, -0.369) | <0.001 | -0.584<br>(-0.874, -0.293) | <0.001 |
| MIMoSA | -0.155<br>(-0.416, 0.105) | 0.239 | -0.191<br>(-0.442, 0.06) | 0.134 |
| WMH-SynthSeg | -1.393<br>(-1.94, -0.847) | <0.001 | -1.278<br>(-1.878, -0.677) | <0.001 |
| nnU-Net-FL | -1.003<br>(-1.479, -0.527) | <0.001 | -0.896<br>(-1.377, -0.416) | <0.001 |
| nnU-Net-FL/T1w | -0.99<br>(-1.368, -0.613) | <0.001 | -0.906<br>(-1.293, -0.519) | <0.001 |
| PLAn-FL | -0.822<br>(-1.157, -0.487) | <0.001 | -0.743<br>(-1.089, -0.397) | <0.001 |
| PLAn-FL/T1w | -0.82<br>(-1.13, -0.51) | <0.001 | -0.749<br>(-1.072, -0.426) | <0.001 |

Age-adjusted models showed similar relationships between WML volumes and clinical disability scores. HF reference WML volumes remained associated with EDSS (β = 0.06 mL^-1^, 95% CI [0.012, 0.109], p = 0.016) and SNRS scores (β = −0.615 mL^-1^, 95% CI [−0.955, −0.274], p < 0.001), as did pULF reference volumes (EDSS: β = 0.051 mL^-1^, 95% CI [0.008, 0.093], p = 0.02; SNRS: β = −0.584 mL^-1^, 95% CI [−0.874, −0.293], p < 0.001). All automated segmentation methods except MIMoSA recapitulated these relationships. Among automated approaches, PLAn-FL and PLAn-FL/T1w showed effect sizes most similar to the HF reference models. No other clinical scores showed significant relationships with segmentation volumes for either reference or automatic segmentation methods, except for 9HPT time associations with WMH-SynthSeg, both nnU-Net models, and both PLAn models before and after age-adjustment. Additional fitted model details are provided in the **Supplementary Materials**.

## 4. Discussion

In this study, we demonstrate accurate automated WML segmentation and clinically relevant WML quantification from pULF-MRI in MS using DL models (nnU-Net and PLAn). PLAn, an nnU-Net-based transfer learning model, showed the strongest segmentation performance across quantitative and qualitative metrics. Furthermore, we show that total WML volume derived from automated segmentation methods tailored for pULF data — WMH-SynthSeg^19^, nnU-Net^20^ and PLAn models — are associated with clinical outcomes comparable to associations from HF WML volume estimates.

These associations highlight the clinical relevance of accurate pULF automatic segmentation in MS, particularly in situations where HF-MRI may not be feasible.

We first assessed WML identification and segmentation at pULF relative to HF-MRI. pULF reference segmentations overestimated lesion volume, likely due to the larger voxel size at pULF causing partial volume effects at lesion boundaries. Even when confluent WML were excluded, 80% of lesions >50 µL (corresponding to a lesion having at least one dimension >4.5 mm) at HF were included in pULF reference. However, a considerable proportion of WML <50 µL, in line with previous studies ^5, 11^, were missed at pULF.

Despite pULF-MRI not capturing smaller lesions seen at HF-MRI, volumes from manual reference segmentations as well as all the automated segmentation models were still significantly associated with clinical measures. This may suggest that pULF visualization of larger lesions is sufficient to capture clinically relevant MS lesions. This aligns in part with the McDonald criteria,^24^ which requires identified lesions to be at least 3mm in one dimension. Additionally, strong volumetric correlations between HF WML segmentation and automated pULF WML segmentation methods in our cohort raises the possibility that pULF may be sufficient to create a representative estimate of total WML burden even when the smaller lesions are not captured. While including smaller lesions in these estimates may be a higher priority in clinical practice, our findings highlight pULF-MRI’s possible utility in clinical trials, potentially allowing for increased MRI data with reduced patient burden, especially for patients with advanced disability who may find standard MRIs challenging to complete comfortably.

Among the algorithms tested, PLAn models exhibited the highest quantitative and qualitative segmentation metrics, demonstrating the most accuracy. PLAn outperformed nnU-Net models, which were more conservative but still outperformed WMH-SynthSeg and MIMOSA. Interestingly, the addition of a different image contrast (T1w) did not improve the performance of either model. This result is specific to pULF, as multi-contrast imaging is typically beneficial for WML segmentation at high field strengths^33, 34^, and suggests that T1w contrast at pULF does not provide additional information for WML. This finding is particularly relevant as it highlights that T2-FLAIR alone can provide sufficient information for accurate WML segmentation without the need for additional scanning, leading to shorter acquisition times for WML burden quantification purposes.

MIMoSA and WMH-SynthSeg achieved the lowest segmentation performance. Their low DSC scores are not unexpected, as these algorithms were applied in an "out-of-the-box" fashion without dataset-specific adaptation, unlike PLAN and nnU-Net. MIMoSA, originally trained on 3T data, had the lowest metrics — an expected result given that pULF-trained models generally outperform those developed for HF MRI. WMH-SynthSeg has previously shown good performance for WML segmentation, but it was not specifically developed for MS lesions. In our dataset, it performed better than MIMoSA but still substantially below models tailored to the pULF domain. Together, these findings suggest that segmentation performance may be limited when algorithms are applied without adaptation to either the target imaging modality or disease context and support the use of pULF-specific training datasets for WML segmentation in MS^5, 19^.

This study has limitations. The pULF dataset used in this study was heterogeneous due to software updates by the manufacturer that effects acquisition parameters, which may have influenced segmentation outcomes. Given the retrospective nature of this study, we included all available cross-sectional data, regardless of differences in software versions, reconstruction methods, or voxel sizes. We did not consider acquisition parameters in the training dataset, despite their potential impact on model performance. Additionally, this is a single site study. Therefore, future work should explore the generalizability of these models in multi-center pULF datasets for further validation.

In conclusion, our findings demonstrate that automated WML segmentation algorithms optimized for pULF MRI not only accurately detect WML but also reflect clinical disability. Reliable WML burden assessment is essential in both MS research and clinical care, offering key insights into disease activity and progression. This is particularly important for clinical trials, where quantitative WML burden is a frequent (often secondary but objective) outcome for interventions^35, 36^. pULF-MRI could serve as an objective tool for evaluating intervention outcomes in clinical trials while minimizing patient burden and allowing for higher number of assessments that can strengthen the statistical power. With its portability and lower cost, pULF-MRI holds great potential to expand access to MRI for MS research.

## DECLARATIONS

**CRediT authorship contribution statement:** Ashley A. Thommana: Conceptualization, Methodology, Data curation, Validation, Formal analysis, Visualization, Writing – Original Draft, Writing – Review & Editing. Corinne A. Donnay: Conceptualization, Methodology, Data curation, Software, Formal analysis, Visualization, Writing – Original Draft, Writing – Review & Editing. Gina Norato: Formal analysis, Writing – Review & Editing. María I. Gaitán: Methodology, Writing – Review & Editing. Ludovica Griffanti: Methodology, Writing – Review & Editing. Govind Nair: Conceptualization, Data curation, Software, Methodology, Writing – Review & Editing. Daniel S. Reich: Conceptualization, Methodology, Writing – Review & Editing, Funding acquisition, Supervision. Serhat V. Okar: Conceptualization, Methodology, Data Curation, Validation, Writing – Review & Editing, Supervision.

## Availability of data and material

Supplemental data, including deidentified clinical data, deidentified imaging data, and deep-learning models (nnU-Net-FL, nnU-Net-FL/T1w, PLAn-FL, and PLAn- FL/T1w), will be released upon publication.

## Data Availability

Supplemental data, including deidentified clinical data, deidentified imaging data, and deep-learning models (nnU-Net-FL, nnU-Net-FL/T1w, PLAn-FL, and PLAn-FL/T1w), will be released upon publication.

## Acknowledgments

We would like to acknowledge and thank Henry Dieckhaus for developing PLAn. We would like to thank the patients and staff of the NINDS Neuroimmunology Branch for their contributions.

---

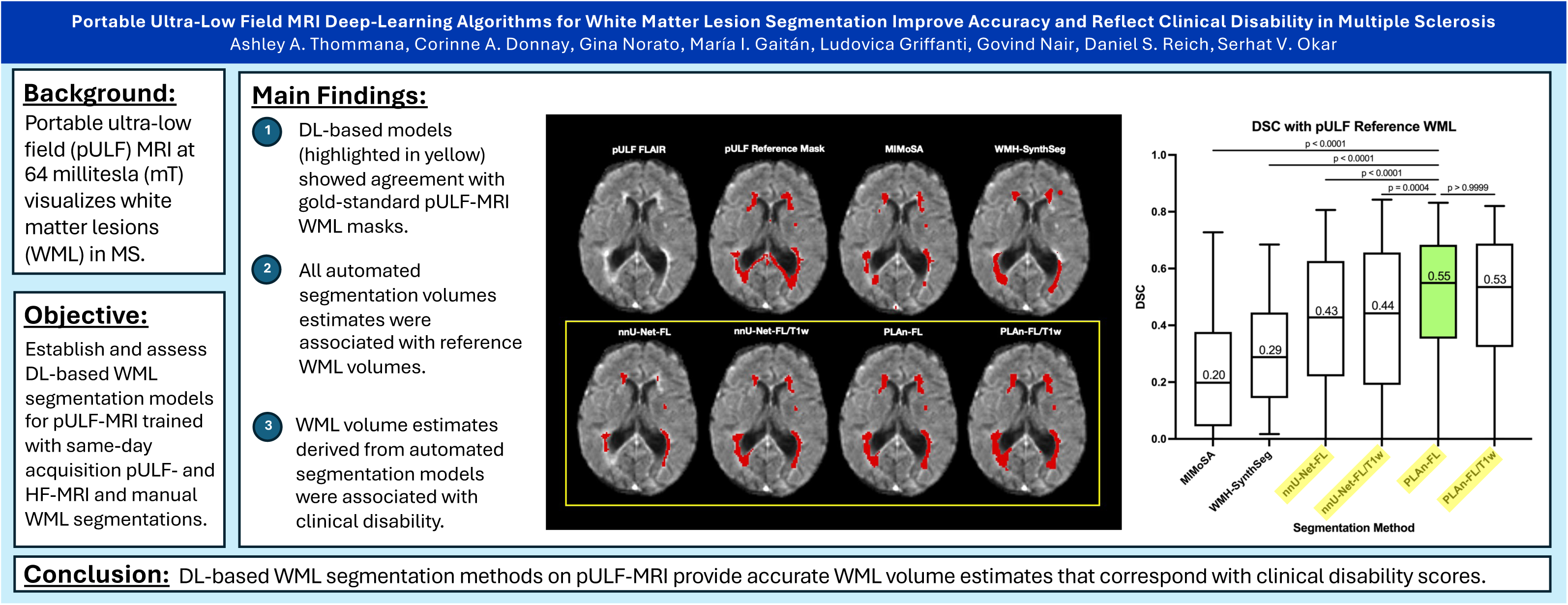

