## Supplemental Materials for "Portable Ultra-Low Field MRI Deep-Learning Algorithms for White Matter Lesion Segmentation Improve Accuracy and Reflect Clinical Disability in Multiple Sclerosis"

**S1. pULF-MRI acquisition details.**

All pULF-MRI data were acquired on the Hyperfine Swoop system at 64 mT. **Supplemental Table 1** provides an overview of acquisition parameters across different software version updates, grouped by acquisition settings.

**Supplemental Table 1: Grouped pULF MRI acquisition parameters showing total number of acquisitions per group and training/testing split.** Legend: TR = repetition time; TE = echo time; TI = inversion time; Ax = Axial; Recon = Reconstruction Method; Soft. Ver. = Software Version; DL = Deep Learning; cart = Cartesian; Part. = Participants; n = number

| **MRI**  **Sequence** | **TR**  *(s)* | **TE**  *(s)* | **TI**  *(s)* | **Flip**  *(°)* | **Recon.** | **Soft. Ver.** | **Slice thickness** *(mm)* | **In-plane voxel size** *(mm^2^)* | **Part.**  *(n)* | **Training/ Testing**  **Split** |
| --- | --- | --- | --- | --- | --- | --- | --- | --- | --- | --- |
| 64mT  3D T2-FLAIR (Ax.) | 3.5 | 0.16 | 1.3 | 90 | cart | rc8.7.0_Beta1 | 5 | 1.6 | 1 | 0/1 |
|  | 3.5 | 0.16 | 1.3 | 90 | cart | rc8.7.0Beta1 | 5 | 1.7 | 48 | 9/39 |
|  | 4 | 0.18 | 1.4 | 90 | cart | 8.3.0 | 5 | 1.8 | 4 | 1/3 |
|  | 4 | 0.18 | 1.4 | 90 | cart | 8.3.2 | 5 | 1.8 | 4 | 1/3 |
|  | 4 | 0.18 | 1.4 | 90 | cart | rc8.3.2 | 5 | 1.8 | 3 | 1/2 |
|  | 4 | 0.19 | 1.4 | 90 | cart | rc8.3.2 | 5 | 1.6 | 1 | 0/1 |
|  | 4 | 0.2 | 1.2 | 90 | cart | rc8.5.0 | 5 | 1.8 | 4 | 2/2 |
|  | 4 | 0.21 | 1.4 | 90 | cart | rc8.5.0 | 5 | 1.6 | 4 | 1/3 |
|  | 4 | 0.14 | 1.4 | 90 | cart | rc8.6.0 | 6 | 2 | 1 | 0/1 |
|  | 4 | 0.17 | 1.4 | 90 | cart | rc8.6.0 | 5 | 1.7 | 12 | 2/10 |
|  | 4 | 0.21 | 1.4 | 90 | cart | rc8.7.0Beta1 | 5 | 1.6 | 1 | 0/1 |
|  | 4 | 0.19 | 1.4 | 90 | DL | 8.2.0 | 5 | 1.6 | 4 | 0/4 |
|  | 4 | 0.19 | 1.4 | 90 | DL | гc8.2.0 | 5 | 1.6 | 13 | 3/10 |
|  | 4 | 0.18 | 1.4 | 90 | DL | rc8.3.0 | 5 | 1.8 | 3 | 0/3 |
|  | 4 | 0.2 | 1.4 | 90 | DL | rc8.3.0 | 5 | 1.6 | 1 | 0/1 |
| 64mT  3D T1w  (Ax.) | 0.9 | 0.006 | 0.4 | 90 | cart | rc8.5.0 | 5 | 1.6 | 1 | 0/1 |
|  | 0.9 | 0.005 | 0.3 | 90 | cart | rc8.7.0Beta1 | 5 | 1.6 | 47 | 8/39 |
|  | 1 | 0.006 | 0.3 | 90 | cart | rc8.6.0 | 5 | 1.6 | 1 | 0/1 |
|  | 1.5 | 0.005 | 0.3 | 90 | cart | 8.3.0 | 5 | 1.6 | 4 | 1/3 |
|  | 1.5 | 0.005 | 0.3 | 90 | cart | 8.3.2 | 5 | 1.6 | 4 | 1/3 |
|  | 1.5 | 0.005 | 0.3 | 90 | cart | rc8.3.2 | 5 | 1.6 | 4 | 1/3 |
|  | 1.5 | 0.005 | 0.3 | 90 | cart | rc8.5.0 | 5 | 1.6 | 7 | 3/4 |
|  | 1.5 | 0.005 | 0.3 | 90 | cart | rc8.6.0 | 5 | 1.6 | 12 | 2/10 |
|  | 1.5 | 0.005 | 0.3 | 90 | cart | rc8.7.0Beta1 | 5 | 1.6 | 3 | 1/2 |
|  | 1.5 | 0.006 | 0.3 | 90 | DL | 8.2.0 | 5 | 1.6 | 4 | 0/4 |
|  | 1.5 | 0.006 | 0.3 | 90 | DL | rc8.2.0 | 5 | 1.6 | 13 | 3/10 |
|  | 1.5 | 0.005 | 0.3 | 90 | DL | rc8.3.0 | 5 | 1.6 | 4 | 0/4 |

**S2. Implementation of MIMoSA and WMH-SynthSeg algorithms**

S2.1 MIMoSA Implementation: To create pULF MIMoSA WML segmentations, preprocessed and skull-stripped pULF FLAIR and registered pULF T1w images in the testing cohort were normalized using the WhiteStripe method^26^. The normalized images were then processed using a pre-trained MIMoSA model that had been originally trained on HF T2-FLAIR and T1w images as described in the original publication⁵. MIMoSA generates probability maps indicating the likelihood of WML presence at each voxel. These probability maps were thresholded at the default value of 0.2 and binarized to produce the final 64mT MIMoSA WML masks.

S2.2 WMH-SynthSeg Implementation: pULF WMH-SynthSeg WML segmentations were obtained by applying FreeSurfer’s “WMH-SynthSeg” tool^18^ to the preprocessed pULF T2-FLAIR images. The WML label was extracted from the automatic segmentation outputs and resampled to the native pULF resolution. WMH-SynthSeg performs simultaneous segmentation of multiple brain structures including WML and outputs segmentations at 1x1x1mm^3^ resolution. The WML label was extracted from the multi-label segmentation output and resampled to the native pULF resolution. To minimize resampling errors and preserve the original lesion volume, all resampled WML masks were linearly interpolated and thresholded at a value that minimized the difference in volume relative to the original lesion mask volume, selected using iterative thresholds in steps of 0.05.

**S3 Pairwise Statistics for pULF Segmentation Overlap Metrics**

For all automatic pULF WML segmentation algorithms, overlap metrics were evaluated against the pULF reference masks using FSL’s *bianca_overlap_metrics*.

WMH-SynthSeg WML masks, originally in 1×1×1 mm³ space, were downsampled to pULF resolution. To avoid potential disadvantages from downsampling, we repeated overlap analysis using WMH-SynthSeg's original output resolution. For this analysis, reference pULF masks and all other segmentation outputs were upsampled to 1×1×1 mm³. Resampling was performed using the *3dresample* tool from AFNI^26^ with linear interpolation and thresholded to minimize the difference in volume relative to the original lesion mask volume using iterative thresholds in steps of 0.05. **Supplemental Table 2** provides DSC comparisons across segmentation methods in both up- and downsampled data. **Supplemental Table 3** highlights overlap statistics (voxel- and cluster-level FPR, voxel- and cluster-level FNR, DER, and OER) across methods.

**Supplementary Table 2: Pairwise statistics between DSC values between segmentation methods and pULF reference WML masks at the original 64mT resolution (downsampled) and at 1.0 m^3^ isotropic resolution (upsampled).**

| **DSC Comparison Across Segmentation Methods (Downsampled)** | **Rank Sum Difference** | **Z-score** | **Adjusted p-value** |
| --- | --- | --- | --- |
| MIMoSA vs. SynthSeg | -80.50 | 3.402 | 0.0100 |
| MIMoSA vs. nnU-Net-FL | -137.50 | 5.810 | <0.0001 |
| MIMoSA vs. nnU-Net-FL/T1w | -153.50 | 6.487 | <0.0001 |
| MIMoSA vs. PLAn-FL | -253.00 | 10.69 | <0.0001 |
| MIMoSA vs. PLAn-FL/T1w | -236.50 | 9.994 | <0.0001 |
| SynthSeg vs. nnU-Net-FL | -57.00 | 2.409 | 0.2401 |
| SynthSeg vs. nnU-Net-FL/T1w | -73.00 | 3.085 | 0.0306 |
| SynthSeg vs. PLAn-FL | -172.50 | 7.289 | <0.0001 |
| SynthSeg vs. PLAn-FL/T1w | -156.00 | 6.592 | <0.0001 |
| nnU-Net-FL vs. nnU-Net-FL/T1w | -16.00 | 0.6761 | >0.9999 |
| nnU-Net-FL vs. PLAn-FL | -115.50 | 4.881 | <0.0001 |
| nnU-Net-FL vs. PLAn FL + T1 | -99.00 | 4.184 | 0.0004 |
| nnU-Net-FL + T1 vs. PLAn-FL | -99.50 | 4.205 | 0.0004 |
| nnU-Net-FL + T1 vs. PLAn-FL/T1w | -83.00 | 3.507 | 0.0068 |
| PLAn-FL vs. PLAn-FL/T1w | 16.50 | 0.6973 | >0.9999 |
| **DSC Comparison Across Segmentation Methods (Upsampled)** | **Rank Sum Difference** | **Z-score** | **Adjusted p-value** |
| MIMoSA vs. SynthSeg | -69.50 | 2.937 | 0.0497 |
| MIMoSA vs. nnU-Net-FL | -133.00 | 5.620 | <0.0001 |
| MIMoSA vs. nnU-Net-FL/T1w | -151.00 | 6.381 | <0.0001 |
| MIMoSA vs. PLAn-FL | -254.00 | 10.73 | <0.0001 |
| MIMoSA vs. PLAn-FL/T1w | -229.50 | 9.698 | <0.0001 |
| SynthSeg vs. nnU-Net-FL | -63.50 | 2.683 | 0.1093 |
| SynthSeg vs. nnU-Net-FL/T1w | -81.50 | 3.444 | 0.0086 |
| SynthSeg vs. PLAn-FL | -184.50 | 7.797 | <0.0001 |
| SynthSeg vs. PLAn-FL/T1w | -160.00 | 6.761 | <0.0001 |
| nnU-Net-FL vs. nnU-Net-FL/T1w | -18.00 | 0.7606 | >0.9999 |
| nnU-Net-FL vs. PLAn-FL | -121.00 | 5.113 | <0.0001 |
| nnU-Net-FL vs. PLAn FL + T1 | -96.50 | 4.078 | 0.0007 |
| nnU-Net-FL + T1 vs. PLAn-FL | -103.00 | 4.353 | 0.0002 |
| nnU-Net-FL + T1 vs. PLAn-FL/T1w | -78.50 | 3.317 | 0.0136 |
| PLAn-FL vs. PLAn-FL/T1w | 24.50 | 1.035 | >0.9999 |

**Supplementary Table 3: Overlap measures across methods.**

Legend: FPR = false positive ratio, FNR = false negative ratio, DER = detection error rate, OER = outline error rate.

| **Method (Downsampled)** | **Voxel-level FPR**  **(mean±SD)** | **Voxel-level FNR**  **(mean±SD)** | **Cluster-level FPR**  **(mean±SD)** | **Cluster-level FNR**  **(mean±SD)** | **DER**  **(mean±SD)** | **OER**  **(mean±SD)** |
| --- | --- | --- | --- | --- | --- | --- |
| MIMoSA | 0.8195±0.1964 | 0.5682±0.2369 | 0.9058±0.09111 | 0.3458±0.2794 | 1.292±0.5917 | 0.2666±0.2215 |
| WMH-SynthSeg | 0.7174±0.2413 | 0.5748±0.1658 | 0.6605±0.2001 | 0.3278±0.2139 | 0.5767±0.5326 | 0.8562±0.3183 |
| nnU-Net-FL | 0.2666±0.2784 | 0.662±0.2327 | 0.172±0.283 | 0.5257±0.2912 | 0.6438±0.6907 | 0.5628±0.3423 |
| nnU-Net-FL/T1w | 0.271±0.2867 | 0.6607±0.2465 | 0.1513±0.2921 | 0.5456±0.2811 | 0.6739±0.7174 | 0.5394±0.3421 |
| PLAn-FL | 0.3484±0.2516 | 0.5447±0.2422 | 0.2377±0.2776 | 0.4417±0.2635 | 0.5059±0.6228 | 0.5304±0.2367 |
| PLAn-FL/T1w | 0.3547±0.2443 | 0.5578±0.2498 | 0.2574±0.2874 | 0.4467±0.2685 | 0.5205±0.6393 | 0.5464±0.2656 |
| **Method (Upsampled)** | **Voxel-level FPR**  **(mean±SD)** | **Voxel-level FNR**  **(mean±SD)** | **Cluster-level FPR**  **(mean±SD)** | **Cluster-level FNR**  **(mean±SD)** | **DER**  **(mean±SD)** | **OER**  **(mean±SD)** |
| MIMoSA | 0.8165±0.1999 | 0.5636±0.2420 | 0.9056±0.0906 | 0.3410±0.2734 | 1.278±0.6004 | 0.2734±0.2239 |
| WMH-SynthSeg | 0.7240±0.2370 | 0.5852±0.1538 | 0.7254±0.1752 | 0.2931±0.2106 | 0.3913±0.5035 | 1.052±0.3910 |
| nnU-Net-FL | 0.2688±0.2757 | 0.6606±0.2355 | 0.1751±0.2835 | 0.5299±0.2904 | 0.6373±0.6920 | 0.5660±0.3391 |
| nnU-Net-FL/T1w | 0.2711±0.2843 | 0.6581±0.2493 | 0.1622±0.2913 | 0.5502±0.2799 | 0.6668±0.7185 | 0.5410±0.3385 |
| PLAn-FL | 0.3499±0.2471 | 0.5415±0.2459 | 0.2482±0.2772 | 0.4445±0.2620 | 0.4993±0.6234 | 0.5306±0.2370 |
| PLAn-FL/T1w | 0.3546±0.2444 | 0.5542±0.2539 | 0.2630±0.2861 | 0.4516±0.2686 | 0.5138±0.6414 | 0.5458±0.2370 |

Overall, overlap metric trends were consistent between downsampled and upsampled analyses across all methods (Supplementary Tables 2, 3), suggesting that the choice to downsample WMH-SynthSeg outputs rather than upsample the reference masks did not meaningfully affect the results.

**S4 Probability Threshold Optimization for pULF Lesion Segmentation Methods**

S4.1 Optimal Probability Threshold Analysis: As shown in main text Table 2, the MIMoSA, PLAn, and nnU-Net models produce probability maps that estimate the likelihood of each voxel being a lesion. The binarized WML masks used in the primary analysis were derived using the default thresholds for each model: 0.2 for MIMoSA, as indicated in the original paper, and 0.5 for both the PLAn and nnU-Net models. To assess the impact of threshold selection on segmentation performance, we recalculated the overlap metrics for these models over a range of thresholds from 0 to 1, with increments of 0.05.

**Figure S1A-E** shows boxplots of DSC across incremental probability thresholds of 0.05, highlighting the boxplot at the default threshold and the threshold with the highest mean DSC.

For all models, there were no significant differences in DSC scores between the default and optimal thresholds. For MIMoSA and both PLAn models, the optimal threshold was similar to the default threshold. By contrast, both nnU-Net models achieved their highest DSC at a lower threshold of 0.1, resulting in a 0.07 improvement in DSC compared to their default settings. These findings suggest that while default segmentations are generally optimal for all models, nnU-Net's under-segmentation might be partially compensated for by thresholding at a lower probability value.

Using the optimal threshold for each model, we compared DSC scores across methods (χ² = 206.4, p<0.0001, **Figure S1F**). Even with these adjusted thresholds, MIMoSA (median: 0.22 [IQR 0.04 – 0.39]) and WMH-SynthSeg (median: 0.27 [IQR 0.14 – 0.43])were not significantly different from each other (Z = 2.22, p=0.34). MIMoSA continued to perform significantly worse than PLAn-FL (median: 0.56 [IQR 0.38 – 0.68]; Z = 11.0, p < 0.0001) and PLAn-FL/T1 (median: 0.55 [IQR 0.35 – 0.68]; Z = 9.1; p < 0.0001), as did WMH-SynthSeg (Z = 8.7, p < 0.0001; Z = 6.9, p < 0.0001, respectively). However, with optimized thresholds, nnU-Net-FL (median: 0.53 [IQR 0.34 – 0.63])and nnU-Net-FL/T1 (median: 0.53 [IQR 0.29 – 0.63])were no longer significantly different from PLAn-FL (Z = 1.9, p = 0.95; Z = 1.9, p = 0.86, respectively) or PLAn-FL/T1 (Z = 0.021, p > 0.99; Z = 0.021, p > 0.99, respectively).

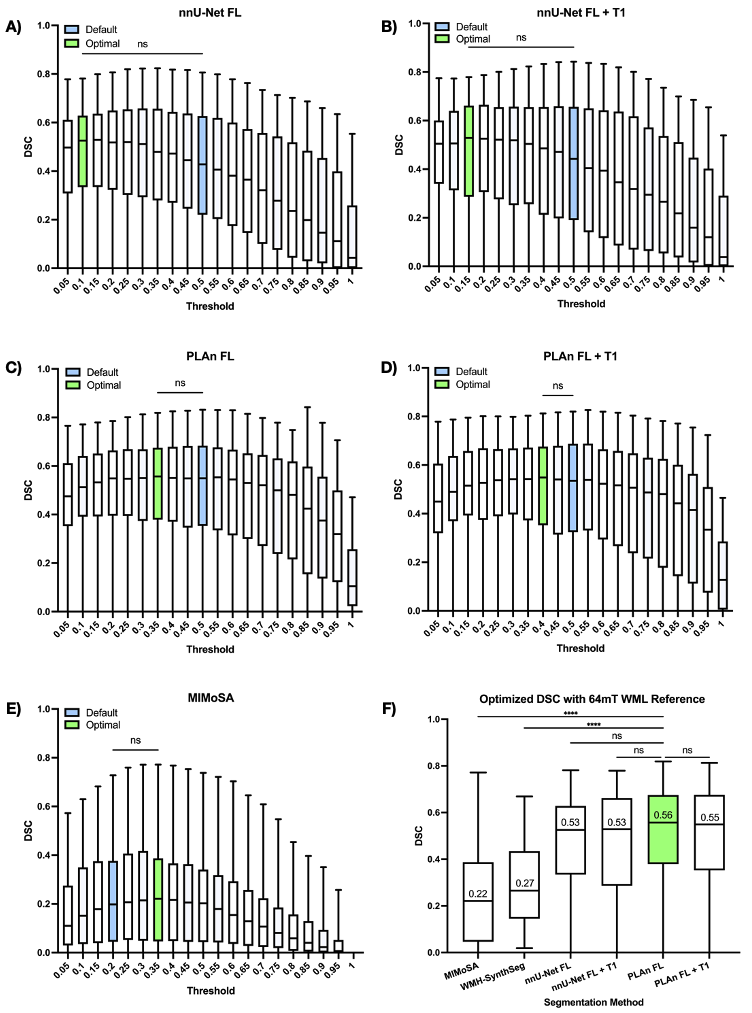

**Supplemental Figure 1:** DSC across different probability map threshold cut-offs for (A) nnU-Net-FL, (B) nnU-Net-FL/T1w, (C) PLAn-FL, (D) PLAn-FL/T1w, and (E) MIMoSA. The DSC at the optimal cut-off for each method (green) was not significantly different than DSC at the default threshold (blue) for any method. (F) Optimized PLAn and nnU-Net models performed similarly to each other and outperformed WMH-SynthSeg and MIMoSA. PLAn FL (green) had the highest mean and median DSC (displayed values).

**S5. Fitted Mixed Linear Model Associations between Clinical Scores and Reference and Automated Segmentation Methods.**

SDMT and PASAT scores were not associated with WML volume estimates produced by any reference or automated segmentation methods. 9HPT times were positively associated with WML volumes from all automated segmentation methods except MIMoSA. They were not significatly associated with reference segmentation WML volumes. These relationships were recapitulated in the age-adjusted models (**Supplementary Table 4)**.

**Supplemental Table 4. Additional associations between WML volume and clinical disability scores across automated matter lesion segmentation methods with and without age-adjustment through fitted multiple linear regression models.**

|  | **Un-adjusted Model** | | **Adjusted Model** | |
| --- | --- | --- | --- | --- |
|  | **β (95% CI) in mL^-1^** | **p-value** | **β (95% CI) in mL^-1^** | **p-value** |
| **SDMT** | | | | |
| HF Reference | -0.44  (-1.104, 0.224) | 0.19 | -0.451  (-1.109, 0.206) | 0.17 |
| pULF Reference | -0.296  (-0.769, 0.176) | 0.21 | -0.281  (-0.751, 0.19) | 0.23 |
| MIMoSA | 0  (-0.407, 0.408) | 0.99 | -0.066  (-0.483, 0.351) | 0.75 |
| WMH-SynthSeg | -0.975  (-2.198, 0.248) | 0.11 | -0.965  (-2.178, 0.248) | 0.12 |
| nnU-Net-FL | -0.34  (-1.118, 0.439) | 0.38 | -0.334  (-1.107, 0.439) | 0.39 |
| nnU-Net-FL/T1w | -0.422  (-1.163, 0.32) | 0.26 | -0.412  (-1.149, 0.324) | 0.26 |
| PLAn-FL | -0.357  (-0.95, 0.237) | 0.23 | -0.358  (-0.947, 0.231) | 0.22 |
| PLAn-FL/T1w | -0.322  (-0.924, 0.28) | 0.29 | -0.322  (-0.919, 0.275) | 0.28 |
| **PASAT** | | | | |
| HF Reference | -0.028  (-0.881, 0.825) | 0.95 | -0.04  (-0.905, 0.826) | 0.93 |
| pULF Reference | 0.146  (-0.291, 0.583) | 0.50 | 0.121  (-0.336, 0.579) | 0.59 |
| MIMoSA | 0.202  (-1.098, 1.502) | 0.75 | 0.203  (-1.116, 1.521) | 0.75 |
| WMH-SynthSeg | 0.008  (-0.82, 0.835) | 0.99 | 0.008  (-0.831, 0.848) | 0.98 |
| nnU-Net-FL | 0.085  (-0.789, 0.959) | 0.84 | 0.084  (-0.802, 0.97) | 0.85 |
| nnU-Net-FL/T1w | 0.008  (-0.82, 0.835) | 0.99 | 0.008  (-0.831, 0.848) | 0.98 |
| PLAn-FL | 0.042  (-0.628, 0.713) | 0.90 | 0.045  (-0.635, 0.725) | 0.89 |
| PLAn-FL/T1w | 0.046  (-0.61, 0.701) | 0.89 | 0.044  (-0.62, 0.709) | 0.89 |
| **9HPT** | | | | |
| HF Reference | 0.504  (-0.083, 1.092) | 0.09 | 0.505  (-0.086, 1.095) | 0.09 |
| pULF Reference | 0.399  (-0.098, 0.896) | 0.11 | 0.413  (-0.085, 0.911) | 0.10 |
| MIMoSA | 0.166  (-0.278, 0.609) | 0.45 | 0.132  (-0.325, 0.59) | 0.56 |
| WMH-SynthSeg | 1.747  (0.638, 2.856)** | 0.003 | 1.765  (0.656, 2.873)** | 0.003 |
| nnU-Net-FL | 0.931  (0.129, 1.732)* | 0.024 | 0.942  (0.139, 1.746)* | 0.023 |
| nnU-Net-FL/T1w | 0.766  (0.081, 1.451)* | 0.029 | 0.783  (0.096, 1.469)* | 0.027 |
| PLAn-FL | 0.616  (0.036, 1.197)* | 0.038 | 0.622  (0.039, 1.204)* | 0.037 |
| PLAn-FL/T1w | 0.6  (0.026, 1.175)* | 0.041 | 0.607  (0.032, 1.183)* | 0.039 |
